# Potential application of Rapid Antigen Diagnostic Tests for the detection of infectious individuals attending mass gatherings – a simulation study

**DOI:** 10.1101/2022.01.02.22268621

**Authors:** Conor G. McAloon, Darren Dahly, Cathal Walsh, Patrick Wall, Breda Smyth, Simon More, Conor Teljeur

## Abstract

Rapid Antigen Diagnostic Tests (**RADTs**) for the detection of SARS-CoV-2 offer advantages in that they are cheaper and faster than currently used PCR tests but have reduced sensitivity and specificity. One potential application of RADTs is to facilitate gatherings of individuals, through testing of attendees at the point of, or immediately prior to entry at a venue. Understanding the baseline risk in the tested population is of particular importance when evaluating the utility of applying diagnostic tests for screening purposes. We used incidence data to estimate the prevalence of infectious individuals in the community at a particular time point and simulated mass gatherings by sampling from a series of age cohorts. Nine different illustrative scenarios were simulated, small (n=100), medium (n=1000) and large (n=10,000) gatherings each with 3 possible age constructs: mostly younger, mostly older or a gathering with equal numbers from each age cohort. For each scenario, we estimated the prevalence of infectious attendees, then simulated the likely number of positive and negative test results, the proportion of cases detected and the corresponding positive and negative predictive values, and the cost per case identified. Our findings suggest that for each detected individual on a given day, there are likely to be 13.8 additional infectious individuals also present in the community. Prevalence of infectious individuals at events was highest with ‘mostly younger’ attendees (1.00%), followed by homogenous age gatherings (0.55%) and lowest with ‘mostly older events’ (0.26%). For small events (100 attendees) the expected number of infectious attendees was less than 1 across all age constructs of attendees. For large events (10,000 attendees) the expected number of infectious attendees ranged from 26 (95% confidence intervals 12 to 45) for mostly older events, to almost 100 (95% confidence intervals 46 to 174) infectious attendees for mostly younger attendees. Given rapid changes in SARS-CoV-2 incidence over time, we developed an RShiny app to allow users to run updated simulations for specific events.

## Introduction

COVID-19 remains a serious threat to public health in Ireland, despite the notably high uptake of vaccination. It thus seems likely that additional nonpharmacological interventions (NPIs) will be necessary to adequately control COVID-19, at least for the near future. However, we would also hope to avoid the more costly NPIs relied on earlier in the pandemic, such as severe limits on movement and gathering, as well as closure of non-essential businesses, schools, and other valuable activities.

In this context, the wider use of rapid antigen diagnostic tests (RADTs) might be useful for helping to control the spread of SARS-CoV-2, and a range of applications have been proposed [1]. The potential value of RADTs follows from the fact that they are less expensive, easier to use, and return results much faster than the diagnostic PCR tests underpinning the Irish COVID-19 surveillance and test-and-trace systems [2]. There is an important trade off however, as RADTs have both lower specificity (Sp) and sensitivity (Se) to the presence of SARS-CoV-2 than PCR diagnostic tests [3], though there is some evidence to suggest that their ability to detect *infectious* cases could be more favourable [4].

Importantly, the performance of RADTs varies by manufacturer [5], and can be strongly influenced by whether the sample is collected by a trained professional vs the person undergoing the test [6], and if the latter, whether the sample is collected is with or without supervision[7], which has knock-on effects for costs. Also, as for any diagnostic test, the performance of RADTs *in practice* will be strongly influenced by prevalence of cases in the tested population. This is because, for a given test sensitivity and specificity, the probability that an individual testing negative is truly uninfected (the negative predictive value, or NPV) will decrease as prevalence increases, while the probability that an individual testing positive is truly infected (the positive predictive value, or PPV) will decrease as prevalence decreases. Given the above, and the fact that the relative costs of false positives vs false negatives are rarely equal, and can change from context to context, it is important that the cost-effectiveness of RADTs be evaluated for each specific use-case.

One potential application of RADTs is to facilitate gatherings of individuals that might otherwise be prohibited, through testing of attendees at the point of, or immediately prior to entry at a venue, previously described as “testing to enable”[1]. Their use in this context has been somewhat evaluated in randomized controlled trials involving RADT based screening of live events (e.g. in Spain [8]). However, such evaluations tended to be conducted when the background prevalence of COVID-19 was relatively low, meaning that the trials were not well-powered to detect impact on SARS-CoV-2 transmission. However, it seems unlikely that similar experiments would be allowed to go forward when the background prevalence is high. Consequently, simulation and/or modelling based approaches might be particularly important for evaluating the use of RADTs in this setting.

The aims of this study were: 1) to estimate the prevalence of infectious individuals within a series of age cohorts by simulating from incidence data and parameters to describe the likely number of infectious days in the population; 2) To simulate mass gatherings with different age-cohort structures to estimate an overall prevalence of infectious individuals; and, 3) to simulate the application of RADTs to these populations to determine the likely utility of these tests at a population level for such events.

## Materials and Methods

### Overall design

With respect to SARS CoV-2, disease occurrence is typically expressed as an incidence reported over a particular time window (e.g. 14-day incidence). However, for the purpose of screening individuals for gatherings, the interest is in the prevalence of infectious individuals at the point in time of the event, rather than the rate at which new infections occur. The prevalence of infectious individuals at any given time point is determined by the incidence of infection as well as the duration of infectiousness for each infected individual. The duration of infectiousness is likely to vary between individuals due to biological variation[9]. In addition, given that public health measures are introduced to identify and limit the ability of infected individuals to transmit the virus, the *number of infectious days in the community* for each infected case will vary depending on whether or not they are detected, at what point in the infectious process they are detected and whether or not they heed public health advice to restrict their movements. Furthermore, some individuals may not present for a test (and are therefore not detected) yet may limit their movements based on self-suspicion. Finally, differences in vaccine use and clinical fraction by age[10] mean that many of these factors, and therefore the anticipated number of infectious days in the community for each infected case, will vary by age cohort.

Consequently, we simulated mass gatherings by sampling from a series of age cohorts. For each age cohort we assumed that the population consisted of five mutually exclusive infected categories (Figure 1), which differed according to the likely number of infectious days in the community for infected individuals in that category. First, *detected* cases were assumed to consist of three groups: 1) Those that were infected and identified through forward tracing by national contact tracing programmes irrespective of their symptom status (Det_CT); 2) those that *were not* identified by contact tracing and were *symptomatic* (Det_Symp), and 3), those that *were not* identified by contact tracing and were *asymptomatic* (Det_Asymp). In addition, it is also recognised that true number of SARS-CoV-2 infections exceeds those that were detected and documented [11]. We assumed that these undetected cases could be considered to comprise of two groups: undetected asymptomatic (Undet_Asymp) or undetected symptomatic (Undet_Symp) cases.

**Figure 1.**
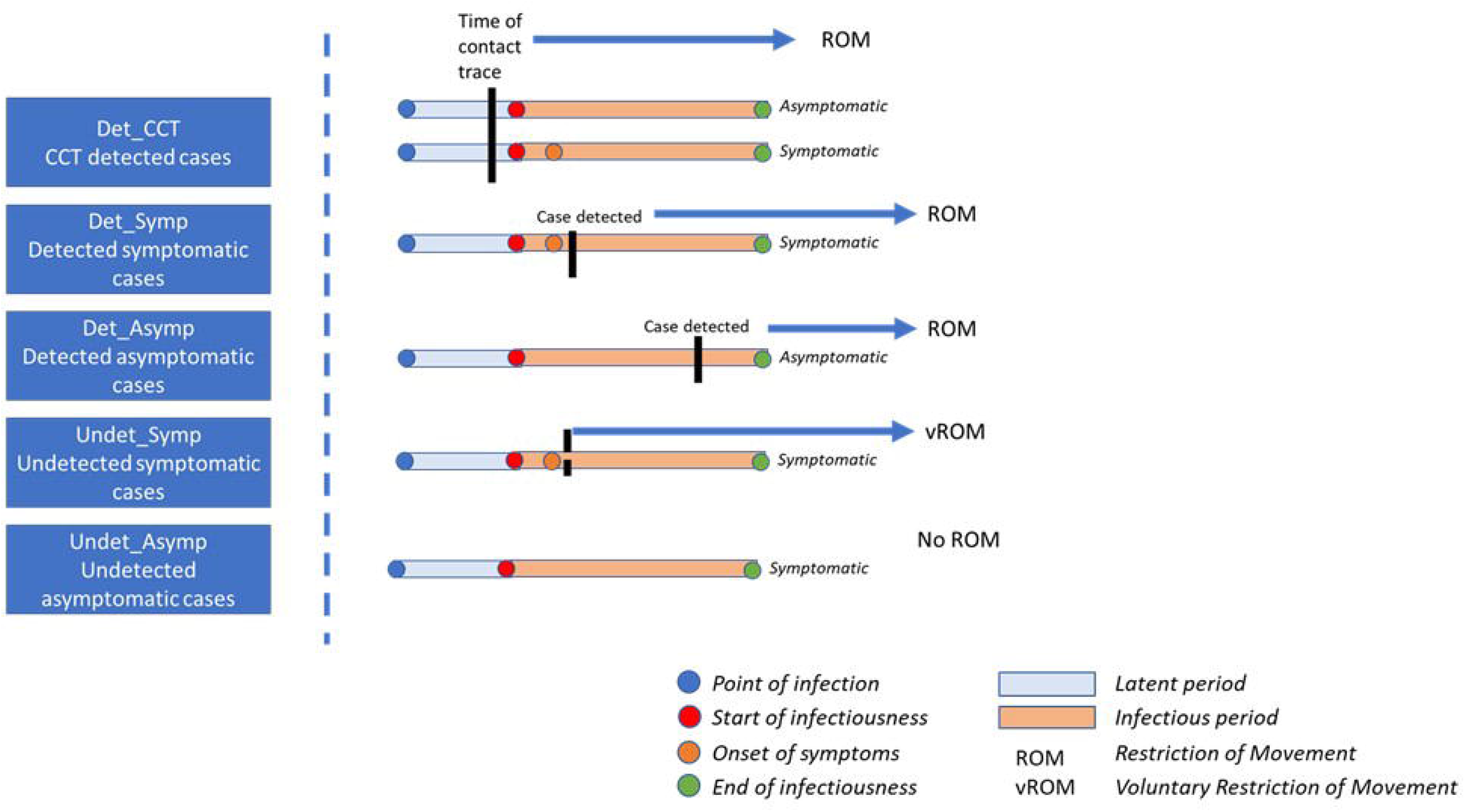
Impact of point of detection for each case category on the number of infectious days in the community.

We used data from contact tracing to estimate the proportion of detected cases in each cohort. Then using probability distributions to represent uncertainty and variability in key parameters determining the number of infectious days in the community for each cohort, we simulated the likely prevalence of infectious individuals for a given 14-day incidence within each age cohort. Gatherings were then simulated with different age cohort structures to estimate the overall prevalence of infectious individuals at the event. Finally, using probability distributions to model diagnostic test sensitivity (Se) and specificity (Sp) for RADTs, we simulated the potential outcome of test applications to these populations in terms of the number of true positives and negatives, number of false positives and negatives, positive and negative predictive values and the cost per case identified.

For the purpose of this study, incidence data was taken from Irish case data published on 6^th^ August 2021 [12]. However, we also developed an R Shiny user interface to allow users to update incidence and proportion of attendees from each age cohort.

### Estimating the proportion of individuals in each ‘case category’

The reported case count of SARS-CoV-2 infections reflects the number of individuals in which the virus has been detected. Individuals may be referred for a test due to contact tracing (i.e. after being identified as a close contact of a confirmed case) or for other reasons (e.g., presentation of symptoms, mandatory testing due to travel). In each case, there was a likelihood that the individual may have been infectious for a number of days prior to being test detected. Depending on the circumstances, the individual may or may not have been observing self-isolation or quarantine.

Contact tracing has been used in an attempt to interrupt chains of transmission. In Ireland, information on the most likely source, date of last contact and the date of the onset of symptoms are recorded for each individual identified as infected [13]. Using previous data collected from contact tracing data[14], we calculated the proportion of detected cases that were previously identified as contacts as Det_CT. For the remaining detected cases, we calculated the proportions that had symptoms, or those that were asymptomatic at the time of contact tracing as Det_Symp and Det_Asymp respectively.

Estimates of the proportion of cases that remain undetected vary across different studies. In reality, changes to the intensity of testing resulting in variation in case ascertainment over time and between regions are likely to impact on these estimates. Mahajan et al. (2021) estimated that 35% of infections were likely detected in the US (data until November 2020) [15], whereas other studies from Europe estimated figures of 40% [11]and 52% [16] from Austria and Italy respectively. In Ireland, three serological studies have been conducted to estimate the proportion of undetected cases, with estimates of the fraction of cases detected ranging from 24-56% [17-19]. Those studies are typically intended to determine the fraction of the population that have developed antibodies indicating exposure to the disease. Frequently those studies also collect information on diagnosed illness of symptoms consistent with the disease. By combining data on sero-prevalence and the diagnostic test accuracy of the antibody tests, the proportion of cases that are undetected can be inferred. For this study, data from the SCOPI study[19] were used in conjunction with a published systematic review of the diagnostic test accuracy of SARS-CoV-2 antibody tests[20] to generate an estimated detected fraction of 50%. We assumed that the majority of these undetected cases were asymptomatic (with no ROM), with the remainder being un-notified/undetected symptomatic cases.

### Number of infectious days per infection category

Individuals from each infection category were assumed to have a number of infectious days in the community that was dependent on a number of different factors: point of infection; latent period; pre-symptomatic and symptomatic infectious period for symptomatic individuals, asymptomatic infectious period for asymptomatic individuals; and point at which the individual went into restricted movement (ROM) (Figure 1).

For notified cases identified through contact tracing we had information on the estimated number of days from exposure to being requested to restrict movements. Combined with a distribution for the number of days from exposure to becoming infectious (i.e., the latent period), we estimated the distribution for the number of days such an individual was potentially infectious and not restricting their movements.

For notified cases not identified through contact tracing, we treated symptomatic and asymptomatic cases separately. For symptomatic cases, we assumed that they would present for testing an average of 1 day after symptom onset (95% CI: 0 to 3 days). Thus they would be infectious and not restricting movements for the pre-symptomatic phase and part of the symptomatic phase. For asymptomatic cases, we conservatively assumed that they would be test detected at a random point after the equivalent of the pre-symptomatic infectious period.

For all notified cases it was assumed that some portion would not adhere to restriction of movements or self-isolation. Compliance with public health guidance on restriction of movement and self-isolation was assumed to be 90% in close contacts and test detected asymptomatic individuals, and 95% in test detected symptomatic individuals.

For undetected cases, it was assumed that asymptomatic cases would not restrict their movements at any point. For undetected symptomatic cases, it was assumed that they would begin to restrict movements at a random point after symptom onset and prior to resolution of symptoms.

#### Det_CCT

The number of infectious days in the population for each case in the Det_CCT category was the period from the end of the latent period up to the point at which the contact tracing phone call was carried out to inform them they were a close contact of a confirmed case. For infected individuals in this category, the time delay was drawn from a distribution fitted to the data of the duration between the last contact with a confirmed case from the Irish national contact tracing data, and the time that CCT were in touch with the contact to inform them of their close contact status. These individuals were assumed to restrict their movements from that period forward and therefore not having further infectious days in the population (Figure 2).

**Figure 2.**
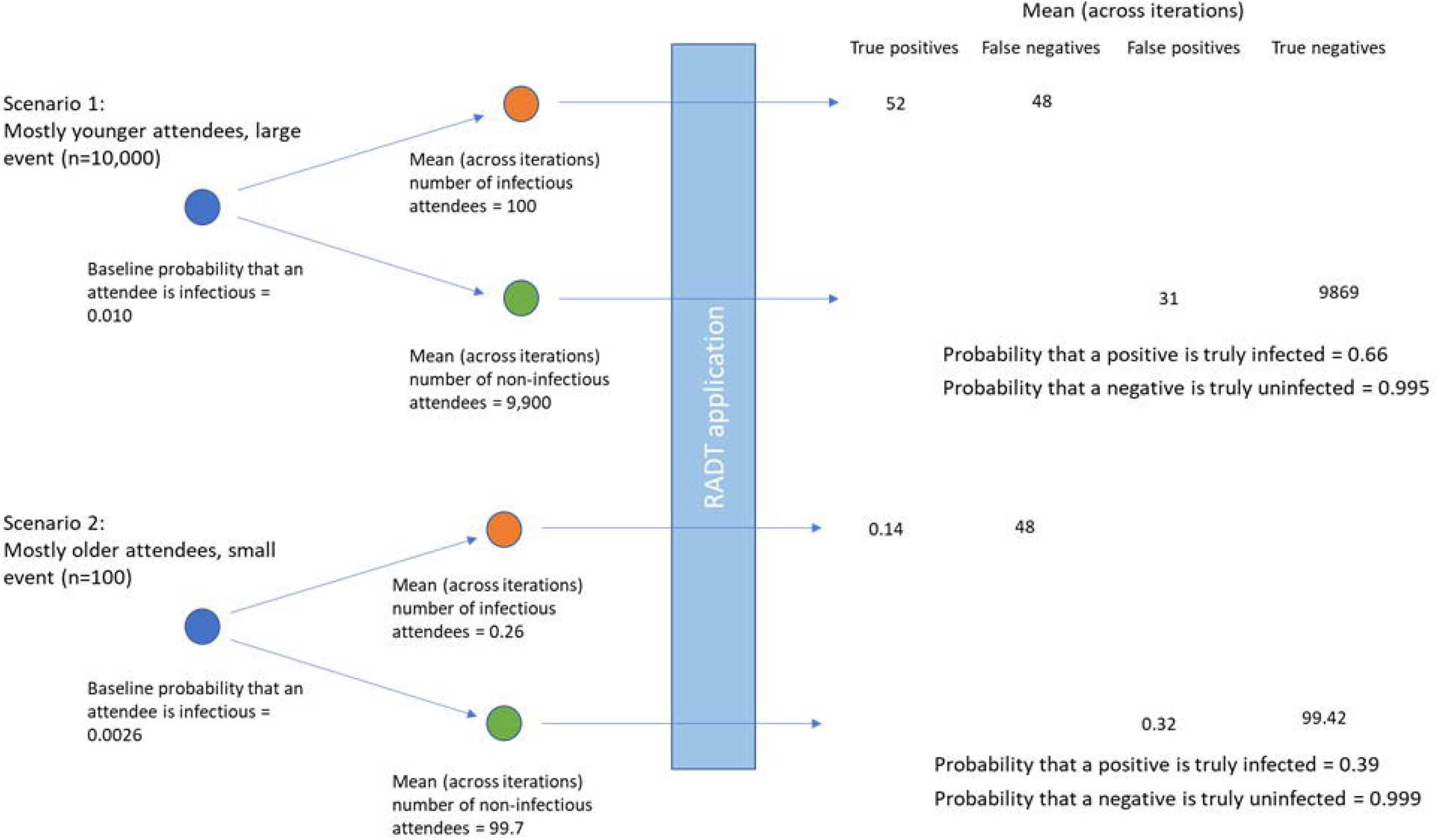
Differences in true and false positives and negatives using two illustrative scenarios.

#### Det_Symp

For those individuals who were symptomatic and detected, they were assumed to restrict their movements shortly after the onset of symptoms. The number of infectious days for this cohort was therefore assumed to be equal to the pre-symptomatic infectious period plus a short duration after the onset of symptoms. For each individual in this cohort, this was simulated using a distribution with a mean of 1 day and a range from 0 to 3 days.

#### Det_Asymp

Individuals that were detected and asymptomatic were assumed to restrict their movements at some point during the infectious period window. These individuals therefore had a number of infectious days that was between 0-100% of the total potential infectious period duration, for each individual in the simulation, this proportion was drawn from a uniform distribution from 0-1.

#### Undet_Symp

Undetected symptomatics were assumed to restrict their movements at some random point after the onset of symptoms. For an individual in this category, they were assumed to have a number of infectious days that was equal to the pre-symptomatic infectious duration plus some fraction of the post-symptomatic infectious period duration. For each individual in this cohort in the simulation, this proportion was drawn from a uniform distribution from 0-1.

#### Undet_Asymp

Those individuals who were undetected and asymptomatic were assumed not to restrict their movement. They were therefore infectious for the total duration of the asymptomatic infectious period. For asymptomatic individuals, the duration of the asymptomatic period was assumed to be the same as the total duration of the infectious period for symptomatic individuals (i.e. the total of the pre and post symptomatic infectious periods).

The distributions used to simulate aspects of the infectious process and number of days infectious are shown in Table 1.

**Table 1.**

| Parameter | Mean | 2.5th – 97.5<sup>th</sup><br>percentile | Distribution<br>(parameter 1,<br>parameter 2) | Source |
| --- | --- | --- | --- | --- |
| Latent period | 3.7 | 1.3 to 8.3 | lognormal (1.2,<br>0.464) | [21, 22] |
| Pre-symptomatic<br>infectious period | 2.4 | 0.4 to 7.9 | lognormal (0.59,<br>0.75) | [22, 23] |
| Post-symptomatic infectious period | 7.1 | 2.8 to 11.5 | Weibull (3.49, 7.90) | [23, 24] |
| Delay from exposure to contact trace | 5.5 | 1.0 to 12.0 | Sampled from observed distribution | Distribution from Irish contact tracing |
| RADT Se | 0.525* | 0.437 to 0.611 | Beta (65.31, 59.18) | [25] |
| RADT Sp | 0.999* | 0.990 to 1.000 | Beta (456.67, 1.46) | [25, 26] |

\*Mode

#### Scenario simulation – estimating the number of infectious individuals per event

We simulated a number of different gathering events. We used 3 different event gathering sizes: 100 individuals, 1,000 individuals and 10,000 individuals. For each, we simulated 3 different age cohorts: 1, ‘Homogenous population’ where equal numbers were drawn from all age cohorts; 2, ‘Mostly younger’ events where 50% of attendees were in the 18-24 years age group, 25% were from 0-18 age group, 25% were from 25-39 age group with no attendees older than 40; ‘Mostly older’ age events where the majority (50%) of attendees were in the 40-65+ age group, 25% were from the 25-39 age group, 25% were older than 65 with no attendees aged 39 or younger.

Using reported incidence, we partitioned cases into each of the categories in Figure 1, and based on the variables in Table 1, created a probability density function for the prevalence within each age cohort. Next, according to size and age makeup of the event, we drew from each of these distributions to generate an overall number of infectious attendees for each scenario, on each iteration. This process was repeated for 10,000 iterations to capture uncertainty and variability in each of the input parameters.

### RADT characteristics

The anticipated number of true positives, false positives, true negatives and false negatives was then calculated by simulating from distributions of sensitivity and specificity of the RADTs. Estimates of the sensitivity and specificity of RADTs for the detection of SARS-CoV-2 infection vary considerably [5, 25]. Much of this variation may be attributed to differences in the definition of the target condition. For example in a large screening study in the UK, the reported test sensitivity changed from 0.40 to 0.91 when the target condition was changed from PCR positive, to PCR positive with a cycle threshold less than 18.3[27].

For the purpose of this study, we assumed that the majority of cases would be asymptomatic and therefore used the asymptomatic subgroup meta-analysis from Brümmer et al. (2021). The sensitivity of the RADT was therefore simulated using a beta distribution with mode of 0.525 and lower 2.5^th^ percentile bound of 0.437[25]. This estimate is similar to a recent report from the use of RADTs in food processing workers in Ireland[26].

Brümmer et al. (2021) reported specificity of RADTs greater than 0.990 across different subgroup analyses[25]. In contrast, a recent study of supervised sampling of asymptomatic individuals in meat processing plants in Ireland reported only 2 false positive results from 5032 samples (0.9996). For the purpose of this study, specificity was simulated using a Beta distribution with a mode of 0.999 and a lower bound 0.990.

To facilitate modification of the proportions of individuals attending the event, we developed an RShiny application allowing decision makers to specify the number of individuals attending the event, the current or likely incidence per age group, the proportion of attendees from each age cohort and to evaluate the impact on the likely number of infectious attendees, number of true and false, positives and negatives.

## Results

Results of the simulation are shown in Table 2. Given the differences in incidence by age group, prevalence of infectious individuals at events was highest with ‘mostly younger’ attendees (1.00%), followed by homogenous age gatherings (0.55%) and lowest with ‘mostly older events’ (0.26%). Consequently, the positive predictive value was lowest in the older aged events (0.39, 95% confidence intervals 0.10, 0.86), and was highest in the younger aged events (0.66, 95% confidence intervals 0.29, 0.96). As an example, Figure 2 shows the difference in numbers of true and false positive and negatives across two illustrative scenarios.

**Table 2.**
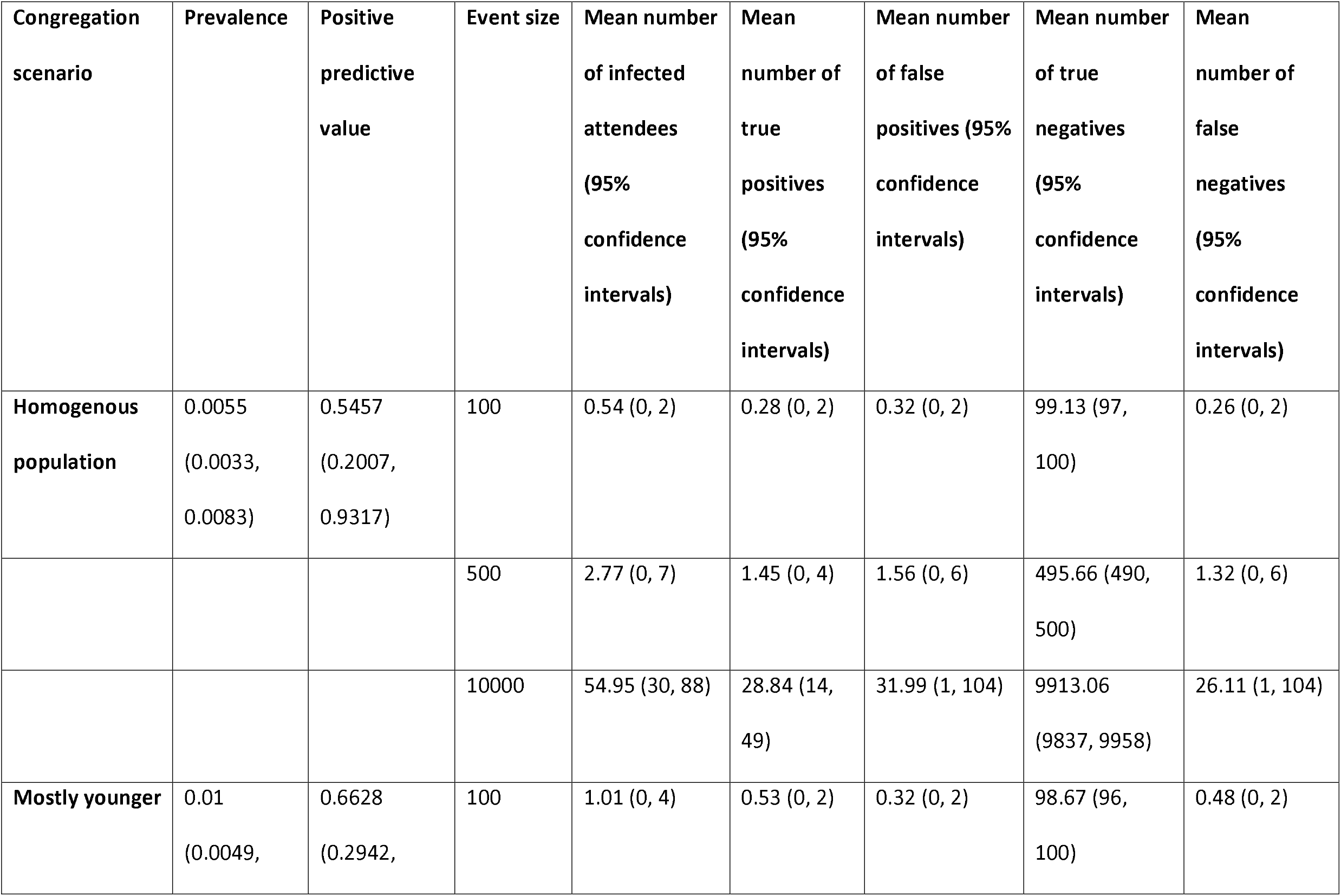
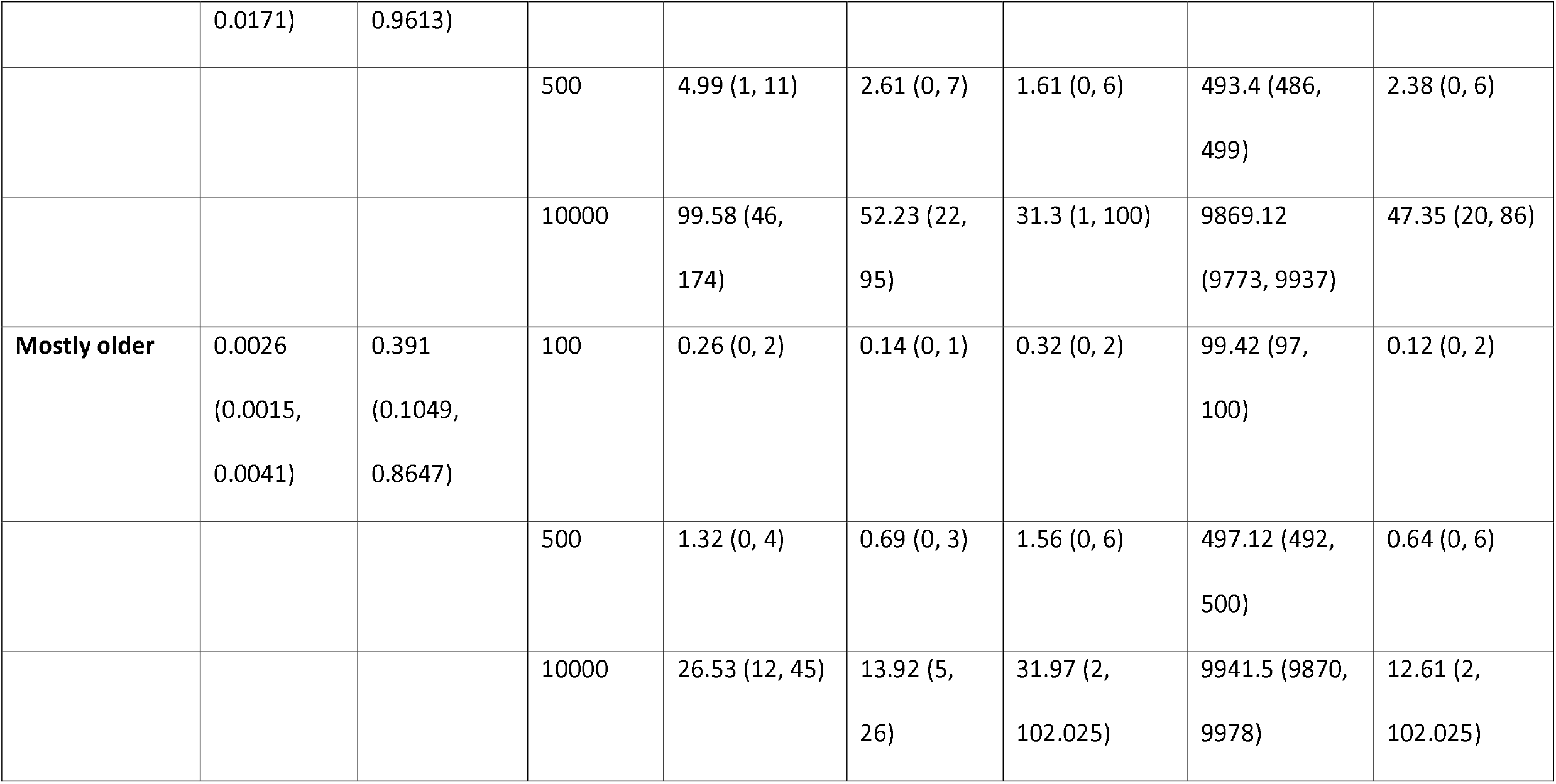

For small events (100 attendees) the expected number of infectious attendees was less than or equal to 1 across all age constructs of attendees. For mostly younger events the expected number of infectious attendees was 1.0 and the 95^th^ percentile was 4, whereas for mostly older events the expected number of infectious attendees was 0.26 with a 95^th^ percentile of 2. For large events (10,000 attendees) the expected number of infectious attendees ranged from 26 (95% confidence intervals 12 to 45) for mostly older events, to almost 100 (95% confidence intervals 46 to 174) infectious attendees for mostly younger attendees.

Given the characteristics of the test, approximately half of these individuals were likely to be detected with the remainder being false negatives. However, there was significant variation with the number of false negatives ranging from 0, 1 for small gatherings of mostly older individuals, to 20 to 86 false negatives at a large gathering of mostly younger individuals.

For homogenous population events the expected number of false positive attendees was 0.3, 1.6 and 31.3 for small, medium, and large events respectively. However, there was also significant variation with this value, with 95% confidence intervals ranging from 0 to 2 for small gatherings of mostly younger attendees, to between 2 and 101 for large gatherings of mostly older individuals.

## Discussion

Understanding the baseline risk in the tested population is of particular importance when evaluating the utility of applying of diagnostic tests for screening purposes. In the context of SARS-CoV-2, case incidence rates are constantly changing. Our study used incidence data to estimate the prevalence of infectious individuals in the community at a particular point in time. However, the methodology can be applied at any time, and therefore has potential as a real-time calculation to support decision making about the control measures required to facilitate mass gatherings while the pandemic is ongoing.

Our findings suggest that for each detected individual on a given day, there are likely to be 13.8 additional infectious individuals also present in the community. This ‘multiplier’ includes individuals who will be detected on subsequent days, as well as asymptomatic individuals who will never be detected. Since undetected asymptomatic individuals do not restrict their movement, they contribute a higher number of infectious days in the community, although these individuals are generally considered to have potentially lower infectiousness than symptomatic individuals[28].

To estimate prevalence from incidence data, we assumed that approximately 50% of cases were detected. This figure is consistent with the international and national literature [11, 15-19]. Undetected cases are a function of numerous factors including testing capacity, disease prevalence, practice regarding referral for testing, and asymptomatic disease. In periods of disease surge, testing capacity comes under pressure and tends to be prioritised for symptomatic cases, increasing the proportion undetected. Frontline healthcare workers are generally highly tested because of their increased risk of exposure and to minimise risk too patients, and as such sero-prevalence studies including healthcare workers may under-estimate the undetected fraction. For this analysis, we used an Irish sero-prevalence study that was conducted during June and July 2020.[19] The study may over-estimate of the undetected fraction on the grounds that it includes infections in the early stages of the epidemic when testing capacity in Ireland was low. Modelling studies have since shown that there was significant under-ascertainment of cases particularly at the peak of the first wave across many countries [29]. Two other Irish sero-prevalence studies have been published: one found a lower undetected fraction (38%) but was focused on hospital-based healthcare workers[18] and a second that was based in primary care and found a higher undetected fraction (73%).[17] For this analysis a wide range of uncertainty was adopted for the undetected fraction to reflect the limited data available and the fact that the undetected fraction is likely to vary over time.

We estimated that the expected prevalence of infectious attendees attending simulated events ranged from 0.2 – 1.0%. This study demonstrates that the prevalence of infectious individuals attending events are likely to be higher for events comprised of mostly younger age cohorts. It is worth noting that the impact of infection in these individuals is much lower [30], however, given contact rates within and between different age cohorts [31], higher rates of infection in in younger age groups is likely to lead to increased community incidence and ultimately, transmission to older, higher risk age cohorts.

Interestingly, our study showed that with older events, the positive predictive value of the test was expected to be less than 40% in all scenarios modelled. Estimates of the specificity of RADT tests are generally very high (in excess of 0.99) [25], however even with ‘higher’ prevalence events made of mostly younger individuals, the absolute prevalence tended to be very low (less than 1%). Given the low prevalence of infectious individuals, the occurrence of false positives although rare, occur at higher frequencies than true positives leading to the low positive predictive value. However, it is worth noting that for this older age cohort, the impact of infection if much higher than for the other age groups simulated[32], therefore a lower positive predictive value might be tolerated.

There are many potential applications of RADT as an aid to the control of SARS-CoV-2 control, Crozier (2021) broadly categorised these as 1) focused asymptomatic testing, 2) focused asymptomatic testing including for example testing for early release from quarantine, and testing to enable otherwise restricted activities, and 3) mass testing. The scenarios modelled in the current study represent only one potential application of the test and should not be used as evidence to support or refute the of RADTs in other contexts [1]. Furthermore, a number of limitations are important to note. For the purpose of this study, we used published estimates of the sensitivity and specificity largely informed by a systematic review and meta-analysis [25]. As is the case for many test validation studies, these values are based on the use of the test relative to a gold standard. For these studies, PCR is used as a pseudo gold standard. However, PCR itself should not be considered a gold standard test and likely has a test sensitivity which is less than 100% [33], therefore the true sensitivity of the RADT may be lower than that used for the analysis. It is also recognised, that ‘reference test’ approaches to diagnostic test evaluation will also likely underestimate the specificity of the evaluated test when the reference test is not a true gold standard [34].

On the other hand, a key consideration for such studies is the target condition which the test aims to identify. In our study, we simulated prevalences of *infectious* individuals by considering the duration of the infectious window and likely restriction of movement of infected individuals. However, this target condition does not necessarily align with the sensitivity estimated in conventional test validation studies which conventionally will include individuals who are PCR detectable but may not be infectious. It has been shown that sensitivity of RADTs varies according to the Ct-value of the corresponding PCR test [26], with transmissibility increasing at lower Ct-values [35]. Estimates of the sensitivity and specificity of any given test will therefore vary according to the distribution of Ct-values within infected individuals in the sampled population. For this study we did not differentiate between different ‘degrees of infectiousness’ for example between detected and undetected infected attendees, or between symptomatic and asymptomatic individuals.

Our analysis also assumed that individuals within age cohorts were independently drawn at random from the overall population. However, it is likely that some degree of clustering is likely to occur with cases, which was not considered in our study. Clustering of cases could result in a disproportionate number of infectious people present at the event, such that realistic range of numbers of infectious individuals potentially attending an event may be greater than those simulated. However, clustering would also imply that those potentially infectious individuals may stay within their grouping and mix less, and therefore it may not lead to markedly increased transmission.

Finally, it is also important to note that the outcomes of the study are conditional on the incidences in a particular region at a particular point in time. In order to facilitate changing underlying incidence, we developed an RShiny app (https://mcaloon-ucd.shinyapps.io/radts/), to estimate the likely number of infectious event attendees, the estimated number of true and false positives and negatives, as well as the positive predictive value and the cost per case identified.

## Conclusions

Understanding the baseline risk in the tested population is of particular importance when evaluating the utility of applying of diagnostic tests for screening purposes, however incidence data underestimate the risk of infectious individuals at a point in time. This study provides a useful method to estimate the likely number of infectious attendees at a particular event. Whilst the disease characteristics are likely to be similar across countries, contact tracing strategies are likely to change within and between countries over time. However with some adaptations, this method could be easily applied across countries.

## Data Availability

All data produced in the present work are contained in the manuscript

